# Estimating seroprevalence of SARS-CoV-2 infection after a highly contagious Omicron outbreak: A cross sectional study in a university setting

**DOI:** 10.1101/2023.08.29.23293775

**Authors:** Ching-Wen Hou, Stacy Williams, Guillermo Trivino-Soto, Veronica Boyle, David Rainford, Selina Vicino, Mitch Magee, Yunro Chung, Joshua LaBaer, Vel Murugan

## Abstract

**Objective:** This study aimed to investigate the seroprevalence of severe acute respiratory syndrome-coronavirus-2 (SARS-CoV-2) antibodies among individuals aged 18 years and older

**Design:** Prospective cohort study.

**Settings:** Population-based study was conducted within the Arizona State University (ASU) community.

**Participants:** The study recruited 1,397 adult participants that volunteered over a period of three days (March 1-March 3 of 2022).

**Primary outcome measures:** Seroprevalence was conducted in the community to assess the presence of SARS-CoV-2-specific antibodies resulting from previous exposure to SARS-CoV-2 and/or vaccination.

**Results:** The seroprevalence of anti-receptor binding domain (RBD) antibodies was found to be 96.3% using a semi-quantitative chemiluminescent immunoassay and 98% using an electrochemiluminescent immunoassay. For anti-nucleocapsid (NC) antibodies, the seroprevalence was 39.1% by an ELISA assay and 41.4% by an electrochemiluminescent immunoassay. Individuals that experienced breakthrough infections exhibited the highest levels of anti-RBD antibodies. Additionally, saliva samples showed promise as a potential diagnostic biofluid for measuring antibody levels, as they exhibited a strong correlation with the data obtained from serum samples.

**Conclusion:** Accurate estimation of population-based serosurveillance for SARS-CoV- 2 will monitor the trend of infection in the community and delineate the geographical spread of the infection. Cumulative incidence of SARS-CoV-2 infection during and after outbreaks is crucial for informing the development of effective risk mitigation protocols within the community. Protocols may include measures such as encouraging booster shots, extending mask mandates, or transitioning to online classes. Serosurveys repeated at regular intervals can also guide containment measures in communities and prompt response to future outbreaks.

**Strengths and limitations of this study:**

- We simultaneously investigated active infection and seroprevalence for the university population.
- Our study was strengthened by having the participants’ self-report data independently validated with diagnostic tests.
- Saliva samples could be a potential diagnostic biofluid for measuring antibody levels.
- Our study was performed within the university setting therefore it only reflects the COVID-19 situation within that community.
- The number of breakthrough infections and the longitudinal samples were small, thus requiring confirmation.

## Introduction

Arizona State University (ASU) is the 6th largest public university in the USA by enrollment, with 79,232 students enrolled during the 2022-23 academic year ^1 2^. It is situated in the southwest region of the USA, with a warm and arid climate, where outdoor activities are counterintuitively more common in the winter than in the summer. In response to the COVID-19 pandemic, ASU switched to remote learning to prevent community-based transmission of SARS-CoV-2 during the first year of the pandemic. However, since the development and distribution of COVID vaccines, the University returned to full in-person learning in Fall 2021.^3^ Understanding whether face-to-face learning has contributed to pathogen spread, especially during the Omicron outbreaks, which is the most contagious variant, is crucial for informing future pandemic response.

ASU has been managing COVID-19 cases since January 2020. It provides a COVID-19 update of known cases in the university community each week. However, most SARS-CoV-2 infections are mild or asymptomatic and may not be detected by surveillance ^4 5^, making the infection-to-case ratio a helpful tool for identifying regions with insufficient testing. Experts estimated that for every virologically confirmed case, about ten infections may have been missed by surveillance systems. Population-based serosurveys are valuable for estimating the proportion of the population previously infected with SARS-CoV-2, providing information about the extent of transmission in the past, and helping to understand the future course of the pandemic ^6–9^.

In 2021, two more transmissible variants of the COVID-19 virus, Delta and Omicron, emerged one after the other. By June 2021, Delta had become the dominant strain worldwide, and it was linked to an uptick in reported school outbreaks. However, in November 2021, Omicron emerged and quickly replaced Delta as the prevailing strain globally by January 2022. The symptoms associated with Omicron infection are generally less severe compared to the Delta, but Omicron exhibits higher transmissibility and shows reduced susceptibility to vaccines. It is important to note that the mortality rate associated with Omicron is lower than that of other strains. ^10–12^ Following a two- and three-month period of sub-strains Delta and Omicron outbreak circulations in ASU, we conducted two population-based serosurveys, one from September 13-17, 2021^13^, and the other from March 1-3, 2022.

The main objective of this study was to measure antibodies against both the spike and NC proteins using various assays to estimate the seroprevalence in the university population after the Omicron outbreak. Our comprehensive analysis of this serosurvey provided critical information: a) The extent of COVID-19 exposure among the university population during the Omicron outbreak; b) the proportion of individuals who have received COVID-19 vaccination and booster doses; and c) the duration of detectable antibodies following vaccination or infection. Furthermore, we compared the findings from this serosurvey with the data obtained during the Delta outbreak serosurvey^13^. This comparison allows us to gain valuable information regarding the infection rate, vaccination rate, and seroconversion rate of these two strains. Additionally, we compared the performance of assays using two different sample types, saliva, and serum, obtained from the same participants. The data collected from saliva samples may have implications for potential use in future serosurveys. Overall, these surveys will help elucidate the trends in antibody response against SARS-CoV-2 proteins during the ongoing pandemic, thereby contributing to our understanding of both natural and vaccine-induced immunity.

## Methods

### Participants

Recruitment for this study was conducted through social media advertising, and potential participants were required to complete a serosurvey before participating in the serosurvey. The recruitment period spanned three days, specifically from March 1 to March 3, 2022, and a total of 1397 participants from ASU were successfully recruited for the study.

### Survey Instruments

Demographic information, COVID-19 vaccination status, testing history, and COVID symptoms were collected through self-reported questionnaire. Participants provided this information voluntarily and will be compensated after giving the blood and saliva samples.

### Blood sample collection

The blood samples were collected by trained phlebotomists at ASU using serum tubes (Cat # 37988 from BD). Within 4 hours of collection, the samples were placed in a cooler for transportation to the clinical testing laboratory at ASU. Upon arrival, the samples were centrifuged at 1300 g for 20 minutes to separate the serum. A total of 1397 serum samples, along with their corresponding survey results, were included in the analysis.

### Saliva sample collection

The saliva samples were collected by participants using saliva collection kits. Prior to collection, participants were instructed to refrain from eating, drinking, smoking, vaping, chewing gum, brushing their teeth, or using oral hygiene products for at least 30 minutes. Participants were advised to wash and dry their hands before the collection process. To collect the saliva sample, participants were provided with a straw and instructed to use it to fill the tube with saliva until it reached the fill lines indicated (excluding bubbles). Once the desired amount of saliva was collected, participants removed the straw and sealed the tube with the provided cap. After sealing the tube, participants were instructed to wipe it with the provided disinfectant wipe and place it in the provided biohazard bag.

### Serology testing

In this survey, serological tests were conducted either at the ASU Biodesign Clinical Testing Laboratory (ABCTL) or the Center for Personalized Diagnostics (CPD). The samples were analyzed for the presence of antibodies against the RBD domain of the Spike protein using the Access SARS-CoV-2 chemiluminescent IgG II and IgM assay (Beckman Coulter) and the Meso Scale Discovery (MSD) coronavirus panel (Meso Scale Diagnostics). These tests were used to estimate the seroprevalence of SARS-CoV-2 antibodies induced by vaccination. Additionally, the samples were tested for antibodies against the NC protein using the Platelia SARS-CoV-2 Total Ab ELISA Assay and the MSD coronavirus panel to estimate the seroprevalence of SARS-CoV-2 infection.

The Access SARS-CoV-2 chemiluminescent IgG II assay from Beckman Coulter is authorized for emergency use and is a semi-quantitative assay. In this study, it was utilized to determine the levels of IgG antibodies against the SARS-CoV-2 RBD protein, following the instructions provided by the manufacturer. ^14^ The assay employed five different concentrations of calibrators and two different concentrations of controls, supplied by the manufacturer, to ensure the integrity of the reagents and proper performance of the assay prior to analyzing the samples. The results obtained were compared to a cutoff value expressed in arbitrary units (AU/mL), which was established during the instrument calibration process.

Access SARS-CoV-2 chemiluminescent IgM assay from Beckman coulter was utilized to measure the IgM antibody level of SARS-coV-2 RBD protein, following the manufacturer’s instructions. ^14^ Two different concentrations of calibrators and controls were provided by the manufacturer and analyzed as part of the assay validation process to ensure reagent integrity and proper assay performance before analyzing samples. The results obtained from the samples were compared to the instrument-defined cutoff value, which is expressed as the signal-to-cutoff (S/Co) ratio.

The Platelia SARS-CoV-2 Total Ab ELISA assay from Bio-Rad is a qualitative diagnostic test used to detect total antibodies (IgM/IgA/IgG) against the SARS-CoV-2 NC protein. The interpretation of the results was based on the guidelines provided by the manufacturer: values < 0.8 were considered negative, values between > 0.8 and < 1.0 were categorized as equivocal, and values ≥1.0 were considered positive.

The Meso Scale Discovery (MSD) coronavirus panel from Meso Scale Diagnostics is a multiplexed immunoassay designed to measure the IgG antibody response to SARS-CoV-2. Each well of the 96-well MSD plate contains different antigens. A calibration curve was established using a reference standard with 4-fold serial dilutions and a zero-calibrator blank for quantitation. The assay also included three levels of controls to ensure the accuracy of the performance. To perform the assay, the plate was first blocked with Blocker A solution for 30 minutes at room temperature (RT). Following three washes with 150 µL/well of MSD wash buffer, 50 µL of calibrator, controls, and diluted samples were dispensed into the plate and incubated with shaking for 2 hours at RT. After incubation and another round of washes, the detection antibody was added and incubated with shaking for 1 hour. Subsequently, the plate was washed with the wash buffer, and reader buffer B was added for reading the plate using the MESO QuickPlex SQ 120 instrument. The multiplexed immunoassay provided quantitative antibody responses to the antigens of interest. The results were reported in AU/mL, as defined during the calibration of the instrument.

### Statistical Analysis

We performed descriptive statistics for demographic variables, vaccination-related variables, and antibody test results. We investigated the relationship between the seroconversion of anti-RBD and anti-NC antibodies and other variables using logistic regressions; and the relationship between anti-RBD antibody level and other variables using linear regressions. Antibody test results between different assays were compared using Venn Diagram analysis and Spearman’s correlation. Antibody decay after post vaccination was plotted and examined using Mann-Whitney test. P-value less than 0.05 was considered statistically significant. R version 4.2.1 and GraphPad Prism 9.5.1were used for statistical analysis.

## Results

### Demographics

The Omicron survey included 1397 participants from ASU that provided both saliva samples for qPCR diagnostic testing and blood donations. Of these participants, 820 (58.7%) were students, 562 (40.2%) were employees, and 15 (1.1%) did not provide information about their occupational status. Among the 1397 participants for whom occupational data were available, 794 (56.8%) were female and 570 (40.8%) were male. Regarding age, 682 participants (48.8%) were 18-25 years of age, 386 (27.6%) were 26-40, 298 (21.3%) were 41-65, 18 (1.3%) were older than 65, and 13 (0.9%) did not report their age. Table 1 presents the demographic characteristics of the different groups.

**Table 1:** Demographics of study participants.

| Variables |  | Serosurvey I* |  |  | Serosurvey II |  |  |
| --- | --- | --- | --- | --- | --- | --- | --- |
|  |  | Students<br>(n=893) | Employees<br>(n=79) | Total<br>(n=1064) | Students<br>(n=820) | Employees<br>(n=562) | Total<br>(n=1397) |
| **Gender** | Female | 444(49.7%) | 51(64.6%) | 556(52.3%) | 409(49.9%) | 381(67.8%) | 794(56.8%) |
|  | Male | 409(45.8%) | 28(35.4%) | 467(43.9%) | 395(48.2%) | 174(31%) | 570(40.8%) |
|  | Other | 10(1.1%) | NA | 11(1.0%) | 16(2%) | 6(1.1%) | 22(1.6%) |
|  | Not Reported | 30(3.4%) | NA | 30(2.8%) | NA | 1(0.2%) | 11(0.8%) |
| **Age** | 18-25 | 723(81%) | 4(5.1%) | 762(71.6%) | 625(76.2%) | 57(10.1%) | 682(48.8%) |
|  | 26-40 | 120(13.4%) | 31(39.2%) | 190(17.9%) | 186(22.7%) | 199(35.4%) | 386(27.6%) |
|  | 41-65 | 19(2.1%) | 44(55.7%) | 81(7.6%) | 7(0.9%) | 289(51.4%) | 298(21.3%) |
|  | >65 | NA | NA | NA | NA | 16(2.8%) | 18(1.3%) |
|  | Not Reported | 31(3.5%) | NA | 31(2.9%) | 2(0.2%) | 1(0.2%) | 13(0.9%) |
| **Race** | White | 410(45.9%) | 62(78.5%) | 528(49.6%) | 272(33.2%) | 380(67.6%) | 656(47%) |
|  | Asian | 270(30.2%) | 6(7.6%) | 292(27.4%) | 331(40.4%) | 62(11%) | 393(28.1%) |
|  | Mixed | 39(4.4%) | 5(6.3%) | 46(4.3%) | 74(9%) | 26(4.6%) | 100(7.2%) |
|  | Black | 24(2.7%) | 1(1.3%) | 27(2.5%) | 22(2.7%) | 15(2.7%) | 37(2.6%) |
|  | Native | 13(1.5%) | NA | 14(1.3%) | 3(0.4%) | 6(1.1%) | 9(0.6%) |
|  | Hispanic or Latino# | NA | NA | NA | 93(11.3%) | 61(10.9%) | 154(11%) |
|  | Other | 99(11.1%) | 4(5.1%) | 117(11%) | 25(3%) | 11(2%) | 37(2.6%) |
|  | Prefer not to say | 7(0.8%) | 1(1.3%) | 9(0.9%) | NA | NA | NA |
|  | Not Reported | 31(3.5%) | NA | 31(2.9%) | NA | 1(0.2%) | 11(0.8%) |
| **Vaccination Status** | Yes | 822(92.1%) | 70(88.6%) | 978(91.9%) | 790(96.3%) | 528(94%) | 1323(94.7%) |
|  | Yes + booster | NA | NA | NA | 453(55.2%) | 423(75.3%) | 879(62.9%) |
|  | No | 67(7.5%) | 9(11.4%) | 82(7.7%) | 29(3.5%) | 32(5.7%) | 61(4.4%) |
|  | Not Reported | 4(0.5%) | NA | 4(0.4%) | 1(0.1%) | 2(0.4%) | 13(0.9%) |
| **Vaccine Source** | Pfizer | 424(47.5%) | 32(40.5%) | 510(47.9%) | 361(44%) | 303(53.9%) | 668(47.8%) |
|  | Moderna | 248(27.8%) | 35(44.3%) | 309(29%) | 242(29.5%) | 201(35.8%) | 443(31.7%) |
|  | Janssen | 86(9.6%) | 2(2.5%) | 94(8.8%) | 74(9%) | 19(3.4%) | 93(6.7%) |
|  | AstraZeneca | 46(5.2%) | NA | 46(4.3%) | 94(11.5%) | 2(0.4%) | 97(6.9%) |
|  | Covaxin | 9(1.0%) | NA | 9(0.9%) | 12(1.5%) | NA | 12(0.9%) |
|  | Sinopharm | 2(0.2%) | NA | 2(0.2%) | 3(0.4%) | NA | 3(0.2%) |
|  | Sinovac | 1(0.1%) | NA | 1(0.1%) | 1(0.1%) | NA | 1(0.1%) |
|  | Not Reported | 77(8.6%) | 1(1.3%) | 93(8.7%) | 33(4%) | 37(6.6%) | 80 (5.7%) |
| **Previously self-reported Covid infection** | Yes | 174(19.5%) | 12(15.2%) | 205(19.3%) | 339(41.3%) | 189(33.6%) | 529(37.9%) |
|  | No | 717(80.3%) | 67(84.8%) | 857(80.6%) | 480(58.5%) | 369(65.7%) | 853(61.1%) |
|  | Not Reported | 2(0.2%) | NA | 2(0.2%) | 1(0.1%) | 4(0.7%) | 15(1.1%) |

\*The data from Serosurvey I were already published in the previous paper<sup>13</sup>

### Self-reported COVID-19 infection and vaccine status

Our study aimed to assess the potential role of asymptomatic carriers in COVID-19 outbreaks in the community, especially during the Omicron outbreak. We investigated the prevalence of PCR positivity in 1397 asymptomatic students and employees from a university community on the days of sample collection and found it to be 0.4% (n=6). Among the 1397 participants, 37.9% (n=529) previously reported testing positive for COVID-19, while 61.1% (n=853) reported no history of a positive test (Table 1).

In the study, 1397 participants were surveyed about their vaccination status. Of these, 879 participants (62.9%) reported being fully vaccinated with a booster, while 1323 participants (94.7%) reported having received at least one dose of the vaccine. Only 61 participants (4.4%) reported never having received a vaccine. Among the vaccinated participants, the majority (47.8%, n=668/1397) received the Pfizer vaccine, followed by Moderna (31.7%, n=443/1397) (Table 1).

### Seroprevalence

#### SARS-CoV-2 RBD of spike IgG

All serological assays were evaluated for detecting anti-RBD IgG antibodies using a set of 1397 serum samples. The results of the study show that the seroprevalence of anti-RBD IgG antibodies was found to be 96.3% and 98% using the Beckman and MSD assays, respectively (Supplementary Table 1). The seroprevalence of anti-RBD IgG antibodies did not differ significantly across the groups, including all participants, students only, and employees only.

Out of the 491 participants who self-reported a history of COVID infection and vaccination (excluding four participants that received attenuated parasite vaccines^2^ and three participants that didn’t provide the source of their vaccine), 488 (99.4%) tested positive for anti-RBD antibodies by Beckman and 484 (98.6%) by MSD for anti-RBD antibody. Among the 30 participants that self-reported a history of COVID infection and were not vaccinated, 21 (70%) tested positive for anti-RBD antibodies by Beckman immunoassay, and 24 (80%) by MSD for anti-RBD antibodies. Of the 820 participants that self-reported no history of COVID infection but were vaccinated, 795 (97%) tested positive for anti-RBD antibodies by Beckman, and 812 (99%) by MSD for anti-RBD antibodies (Supplementary Table 2).

#### SARS-CoV-2 NC antibodies

Overall, the seroprevalence for total anti-NC was 39.1% by Bio-Rad and 41.4% for anti-NC IgG by MSD (Supplementary Table 1). Out of the 529 participants that reported having had COVID-19, regardless of their vaccination status, 381 (72%) tested positive for anti-NC antibodies using the Bio-Rad assay, and 385 (72.8%) tested positive using the MSD assay (Supplementary Table 2). Among the 836 participants that reported no previous history of COVID-19 (excluding 12 participants who received attenuated parasite vaccines, three that did not provide vaccine information, and two that did not report their vaccination status), 17.3% (n=145) and 20.7% (n=173) tested positive for anti-NC antibodies using the ELISA and MSD assays, respectively, suggesting they had had a previous, undetected COVID-19 infection (Supplementary Table 2). These results suggest that there was a substantial number of asymptomatic or mild COVID-19 cases that went undetected, which could have implications for public health measures.

#### Demographic variables and seroconversion

Seroconversion refers to the timepoint when antibodies against a virus are detected in the blood following viral infection or vaccination. In our study, we conducted sub-group analyses to investigate potential associations between seroconversion rates in the ASU community and demographic factors of race, gender, age, employment status, and vaccine types. Our findings indicate that there were no significant differences observed among different races, age groups, gender, and employment status in terms of their ability to generate anti-RBD antibodies following self-reported vaccination. In line with other reports^15 16^, individuals that received mRNA vaccines demonstrated significantly higher seroconversion rates compared to those that received other vaccine types. Interestingly, our analyses also indicated that White participants were less likely to seroconvert anti-NC antibodies following self-reported exposure to the SARS-CoV-2 virus compared to Others (odds ratio (OR)=0.50 (95% confidence interval (CI): (0.28, 0.88) compared to Asian, p=0.02; OR=0.55 (95% CI: (0.33, 0.91), p=0.02) compared to the all other participants other than Asian, p=0.02) (Supplementary Table 3).

Furthermore, our analysis revealed no significant differences in the rate of anti-RBD antibody level among these demographic groups. Notably, participants that received mRNA vaccines, specifically Pfizer or Moderna, exhibited slower antibody decay compared to other vaccine types (beta=129.71, (95% CI: (86.79, 172.63)), p<0.001), Interestingly, participants aged 50 and above showed higher anti-RBD antibody levels than those in the 30-40 age group (beta=67.60, (95% CI: (11.64, 123.56)), p=0.02) or 40-50 age group (beta=85.85, (95% CI: (25.98, 145.71)), p<0.01) (Supplementary Table 4).

Similarly, when analyzing serosurvey I and II together using the same analysis, we observed that White participants had a significantly lower likelihood of seroconverting anti-NC antibodies compared to Asian participants (OR=0.40 (95% CI: (0.24, 0.66)), p<0.001) (Supplementary Table 5). Interestingly, we observed that White participants had lower anti-RBD level compared to non-Asian participants (beta=-34.22, (95% CI: (- 60.90, -7.53)), p=0.01). Additionally, participants in the 40-50 age group exhibited lower anti-RBD level compared to those in < 20 age group (beta=-64.39, (95% CI: (-116.87, - 11.92)), p=0.02) and 20-30 age group (beta=-50.70, (95% CI: (-97.90, -3.50)), p=0.04). Also, participants aged 50 and older showed higher anti-RBD level compared to those 30-40 (beta=55.74 (95% CI: (10.38, 101,10)), p=0.02) and 40-50 (beta=83 (95% CI: (34.38, 131.63)), p<0.001) (Supplementary Table 6). These findings suggest potential variations in immune response based on both race and age within the ASU community using a large group set. However, further investigation is necessary to better understand the underlying factors contributing to these observations.

#### Comparison of assays performances

The Venn diagrams in Figure 1A illustrate the distribution of positive results for two different assays measuring seropositive responses to the RBD of the spike protein. In addition to the 1330 specimens that were positive for both assays, 16 and 39 specimens were exclusively positive for Beckman and MSD, respectively. The percentage of positive results for both assays for anti-RBD IgG were comparable (96.3% and 98%, respectively), as shown in the Supplementary Table 1, which is based on the same sample population. Furthermore, Figure 1C displays the correlation (r=0.89) between the values of anti-RBD IgG measured by the two assays.

**Figure 1.**
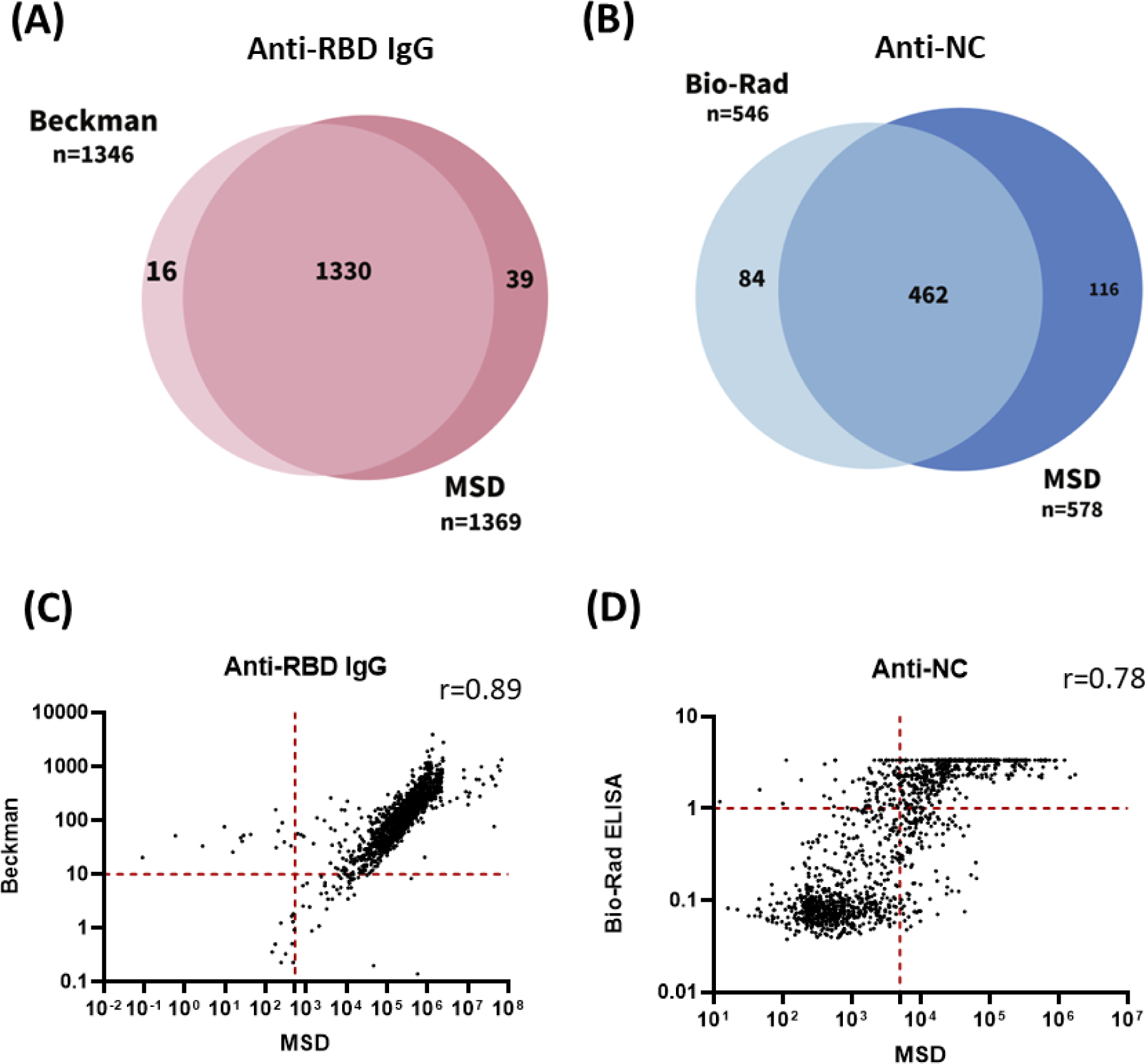
Comparison and correlation of assays. Venn diagrams showing overlap of positive results of **(A)** RBD of Spike and **(B)** Nucleocapsid from two different assays. **(C)** Correlation between the value of anti-RBD IgG by Beckman and the MSD assay. **(D)** Correlation between the value of total anti-NC by Bio-Rad assay and the value of anti-NC IgG by the MSD assay. A red dotted line indicates the cutoff line. All test values equal to or greater than this line were considered positive. r is calculated using Spearman correlation.

Figure 1B shows the overlapping distribution of positive results for two different assays measuring seropositive NC. In addition to the 462 specimens that were positive for both Bio-Rad and MSD assays, 84 and 116 specimens were exclusively positive for Bio-Rad and MSD, respectively. In addition, Figure 1D displays a positive correlation (r=0.78) between the values of anti-NC antibody level measured by Bio-Rad and MSD assays, with somewhat more disagreement than that observed for anti-RBD IgG.

#### Anti-RBD IgG antibody levels after vaccination

Previous studies have demonstrated a decline in anti-SARS-CoV-2 antibody levels during the first six months after COVID vaccination. ^17–19^ In our study, we investigated the persistence of anti-RBD antibody titers among participants that received a COVID vaccination without self-reported prior infection (Figure 2A, indicated in red). Our findings reveal that antibodies could still be detected 12 months after vaccination. Additionally, we also compared these findings with the data obtained from Serosurvey I conducted in September 2021 (shown in blue in Figure 2A). Both serosurveys demonstrated a similar trend of declining anti-RBD antibody levels over time. For Serosurvey I, the median anti-RBD antibody level among participants three months after vaccination was 84.42 AU/mL (interquartile range [IQR], Q3-Q1: 176.51). In Serosurvey II, the corresponding value was 143.97 AU/mL (IQR: 154.045). Notably, the most substantial decline occurred between three months and the 4–6-month post-vaccination period in both Serosurveys (*p*<0.0001 for both). Subsequently, antibody levels continued to decrease, reaching 27.2 AU/mL (IQR: 48.71) and 37.09 AU/mL (IQR: 56.02) for participants 7-9 months after vaccination in Serosurvey I and Serosurvey II, respectively. The median level of anti-RBD antibodies among vaccinated participants in Serosurvey II were significantly higher compared to Serosurvey I in the 0-3 months (143.97 AR/mL (IQR:154.05) vs 84.42 AU/mL (IQR:176.51), *p*<0.0001).

**Figure 2.**
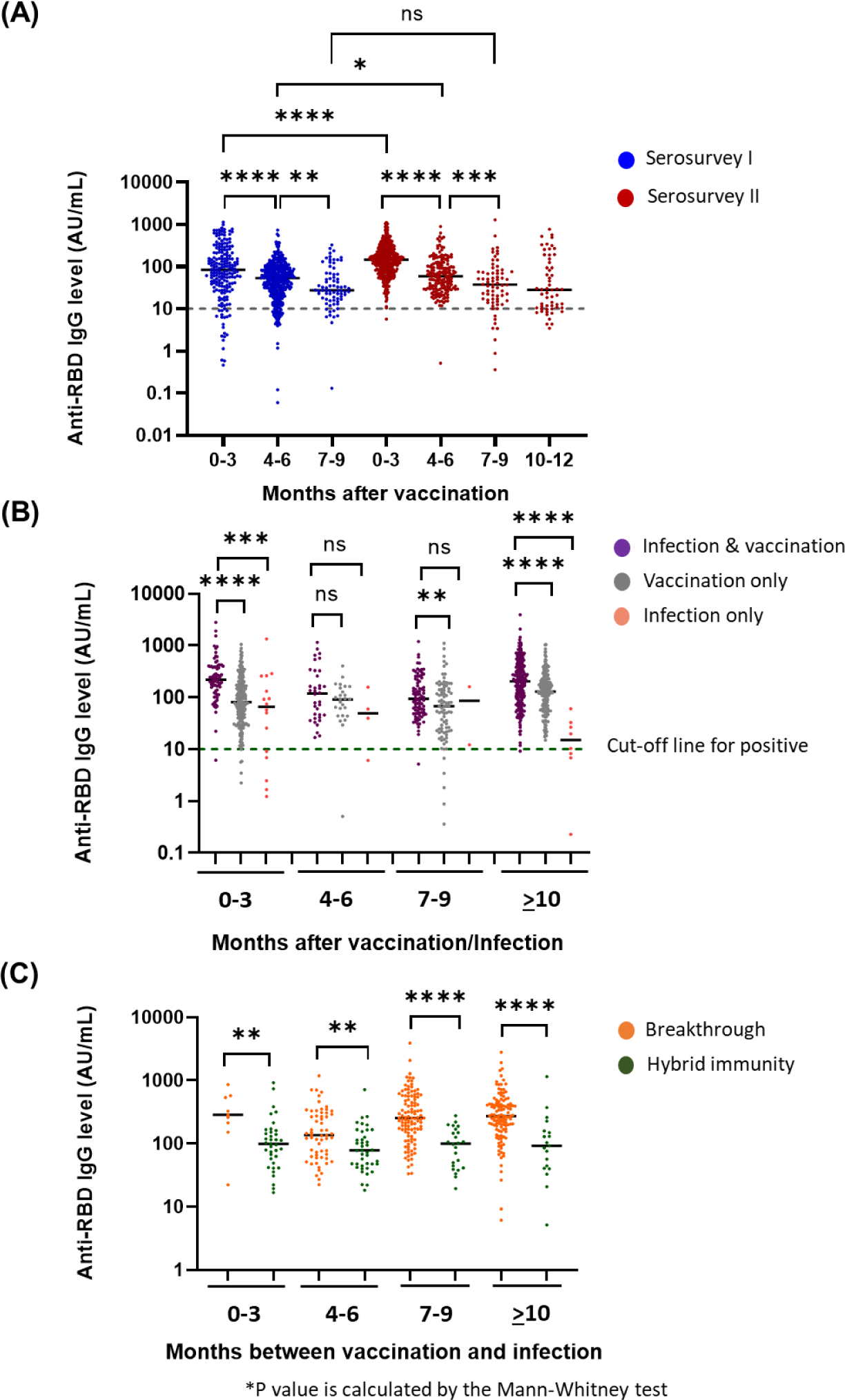
Antibody response in participants with previous COVID infection, vaccination, or both. Anti- RBD antibodies were measured using Beckman immunoassay in **(A)** participants that had previous COVID vaccines without self-reported prior infection, and **(B)** participants that had previous COVID infection or COVID vaccines or both. Participants were categorized by the vaccine or COVID infection and time interval from vaccination/infection to blood sample collection. **(C)** Participants were categorized based on the order and approximate time scale of COVID infection and vaccination for each group. Cutoff defined per manufacturer. *P values are calculated by the Mann-Whitney test. *p-value <0.05, **p-value <0.01, ***p-value <0.001 and ****p-value <0.0001.

#### RBD Antibody responses following vaccination/infection

The study initially divided participants into various categories based on their vaccination and infection status, including those that had received only the vaccine, those that had been infected with COVID-19 only, and those that had experienced both. The participants were then further grouped based on the duration between their most recent vaccination or infection date and the collection date, dividing them into four categories: 0-3 months, 4-6 months, 7-9 months, and ≥ 10 months.

Among all groups, the median level of anti-RBD antibody levels was found to be higher in the subgroups of vaccinated participants that had also been infected with COVID-19 compared to those that had received only the vaccine or had been infected only. Notably, the participants that had never been vaccinated (infection only) had the lowest median anti-RBD antibody level in each group, contrasting with the hybrid group (vaccination and infection) (Figure 2B). Median (interquartile range (IQR)) of anti-RBD antibody levels among participants three months after hybrid (vaccination and infection) and infection only were 220.28 (IQR: 256.30) AU/mL and 65.96 (IQR:154.66) AU/mL respectively. This pattern persisted across other time periods as well. Among participants 4-6 months after the hybrid and infection only, the levels were 118.16 (IQR: 232.48) and 49.57 (IQR: 52.88) AU/mL, respectively. For those 7-9 months after the hybrid and infection only, the levels were 94.25 (IQR: 149.52) AU/mL and 86.33 (IQR: 74.18) AU/mL, respectively. Finally, among participants beyond 10 months after the hybrid and infection only, the levels were 204.93 (IQR: 298.63) AU/mL and 15.07 (IQR: 20.22) AU/mL, respectively.

#### Increased anti-RBD IgG levels after breakthrough infection

The study then examined whether breakthrough COVID-19 infections were associated with an improved immune response. The participants were divided into two groups: those that had experienced infections after vaccination despite being fully vaccinated (vaccine first), and those that had received vaccinations after being infected with SARS-CoV-2 (infection first).

The analysis revealed that the group with breakthrough infections after vaccination (orange dots in the Figure 2C) had significantly higher levels of anti-RBD IgG antibody levels ranging from 1.7 to almost 3 times those of the group with prior infection-induced immunity across various time periods (in green in Figure 2C), showing an association between vaccine first and enhanced immune response.

#### Decreased anti-NC IgG antibody levels after infection

In our study, we examined the antibody response to the SARS-CoV-2 NC protein in individuals that had been previously infected with the virus. However, the ELISA assay (Bio-Rad) we used to detect NC antibodies was qualitative only. Therefore, to obtain quantitative data on the trend of NC antibody levels after infection, the MSD immunoassay from Meso Scale Diagnostics was applied. Participants were stratified into several time intervals (0-3, 4-6, 7-9, 10-12, 13-15, 16-18, and >18 months) based on the duration between their infection and sample collection dates. Similar to RBD antibody levels, NC antibody levels showed a decline over time. However, we observed an increase in the level of NC antibodies after 7-9 months post-infection (Figure 3A). Among the 529 participants in the ASU community that reported a history of COVID infection, 81 participants tested negative on both assays, potentially due to antibody decay following their SARS-CoV-2 exposure (Figure 3B).

**Figure 3.**
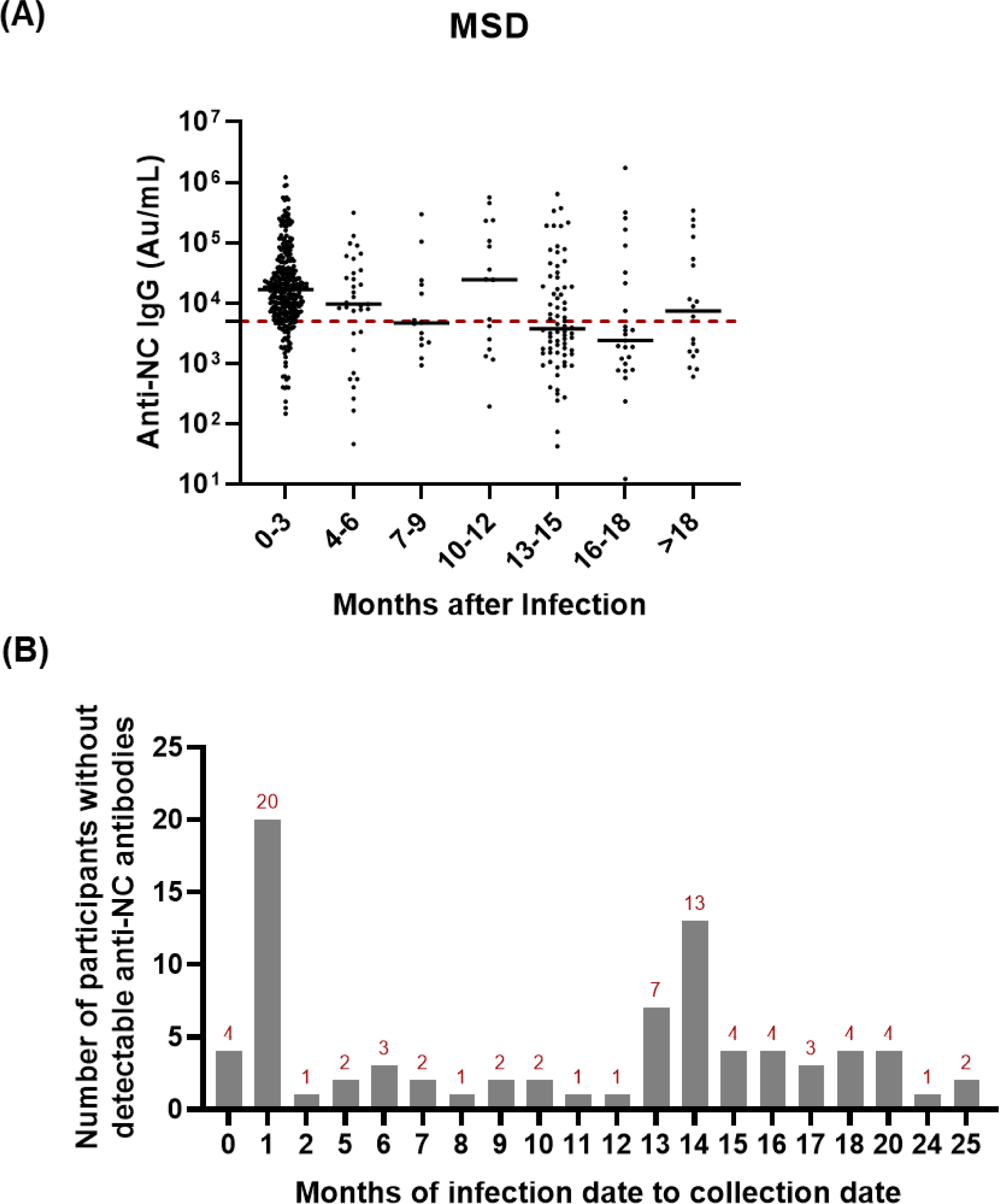
Anti-NC antibody decay post-infection. **(A)** Anti-NC antibodies were measured using MSD. Participants were categorized by the COVID-19 infection and the time interval from infection to blood sample collection. **(B)** Number of participants that had a previous COVID infection and did not exhibit detectable levels of anti-NC antibodies.

#### Comparison of two surveys

Two surveys were conducted with 1,064 and 1,397 participants recruited in September 2021 and March 2022, respectively. Six months after the initial survey, more people were found to be infected with COVID-19 based on both self-reported data and positive results of anti-NC antibodies. Moreover, the proportion of people with detectable anti-RBD antibodies increased from 88% in September 2021 to 96.4% in March 2022 when using the Beckman assay, and from 97% to 98% when utilizing the MSD assay (Supplementary Table 7).

#### Longitudinal samples from two serosurveys

A total of 137 participants, comprising 9.8% of the sample, had also taken part in a previous survey conducted in September 2021^13^. Longitudinal serological analysis of their serum samples revealed an increase in the median levels of both anti-RBD and anti-NC antibodies over the six-month period between the two surveys.

Initially, the 137 participants were categorized based on their self-reported vaccination and infection status prior to serosurvey I, which is listed on the left side of Figure 4. The categories included individuals that had both previous COVID infection and vaccination, those that had only vaccination, those that had only COVID infection, and those that had neither previous COVID infection nor vaccination. Subsequently, the four groups were further categorized into three categories based on their self-reported status before Serosurvey II, which is listed at the top of Figure 4. The categories included individuals who had no infection or vaccination, those who had only infection, and those who had only vaccination (booster) after Serosurvey I. Each graph in Figure 4 represents a before-and-after plot, and each dot represents one participant. Among participants whose status did not change (i.e., they did not get infected with COVID-19 or receive a booster vaccine between surveys) in each group (first column of Figure 4), the level of anti-RBD antibodies decreased. However, we observed that some participants’ anti-RBD antibody levels increased even though they reported no change in their status after Serosurvey I. In contrast, among participants that had either COVID-19 infection or a booster vaccine between the two surveys, the anti-RBD antibody level increased in each group (second and third columns of Figure 4).

**Figure 4:**
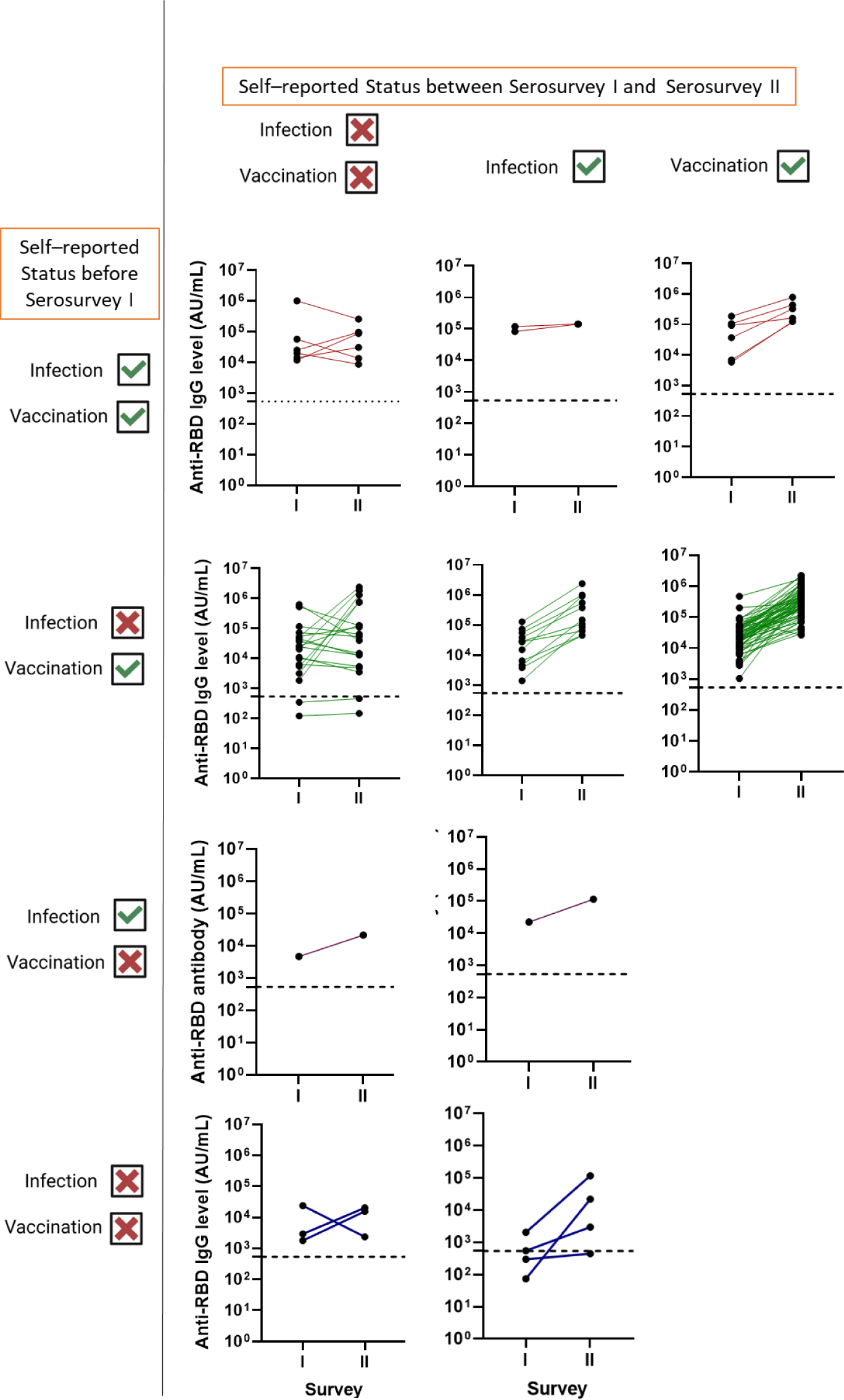
Anti-RBD antibodies in participants that participated in both serosurveys. Common participants were categorized by their status of vaccine and COVID-19 infection between Serosurvey I and II.

#### The levels of anti-RBD antibodies in the saliva correlate with the levels of antibodies in the serum

Saliva collection is a non-invasive method that allows individuals to self-collect samples at home. However, it has received relatively little attention for detecting antibodies to SARS-CoV-2 antigens due to its lower concentration of antibodies compared to serum samples ^20^. In our study, Saliva and serum samples were collected on the same day. In our initial investigation, we examined whether antibody levels to RBD in the saliva correlated with those measured in the serum. First, we selected 20 negative saliva samples from Serosurvey I, considering their self-reported vaccination and COVID infection status, as well as the absence of anti-RBD and anti-NC antibodies, in order to calculate the cutoff value for saliva samples. Next, we found a significant positive correlation (r=0.74) between the levels of anti-RBD antibodies in paired saliva and serum samples (n=1384) (Figure 5A). Furthermore, we conducted a comparison between saliva data from MSD and serum data from an EUA-approved assay (DxI from Beckman). The correlation coefficient between these two data sets was 0.76. Additionally, the Positive Percent Agreement (PPA) and Negative Percent Agreement (NPA) were found to be 96.8% (95.7%, 97.6%) and 51.9% (34%, 69.3%) respectively (Figure 5B). These findings suggest that saliva may represent a viable alternative for antibody testing.

**Figure 5.**
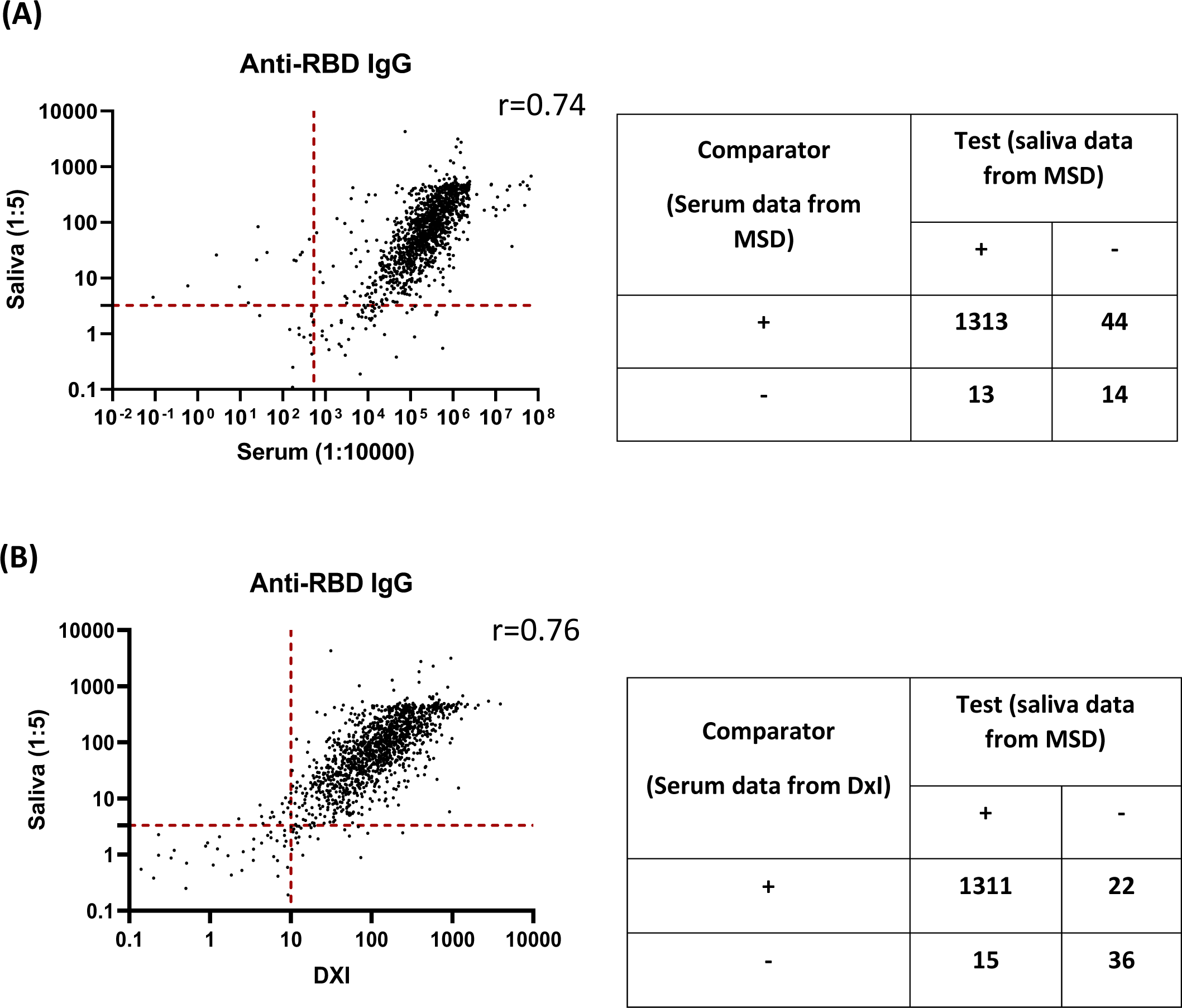
The correlation of sample types and assays. **(A)** Correlation of the value of anti-RBD IgG between serum and saliva samples. **(B)** Correlation between the value of anti-RBD IgG using serum samples by DXI (Beckman) and using saliva samples by the MSD assay. A red dotted line indicates the cutoff line. All test values equal to or greater than this line is considered positive. r is calculated using Spearman correlation.

#### Antibody levels responded to SARS-CoV-2 Omicron subtype variants using saliva samples

To investigate the RBD antibody levels against the SARS-CoV-2 Wuhan wild type, Alpha, Beta, Delta, and various subtypes of Omicron following the Omicron outbreak, the MSD platform was applied. As anticipated, the median level of anti-RBD antibodies against the Wuhan wild type was significantly higher compared to other variants.

Furthermore, we observed a significant decrease in the median antibody levels against each subtype of Omicron variants when compared to the Delta variant (Supplementary Figure 1).

## Discussion

Accurately estimating the cumulative proportion of the population infected with SARS-CoV-2 in a university community during or after an outbreak is crucial for effective planning and targeted public health responses in future outbreaks. This is particularly important considering the limited availability of seroprevalence studies specifically focused on universities. By obtaining these data, we can enhance our understanding of the extent of exposure and immunity within the university setting, enabling us to develop tailored strategies to mitigate the impact of future outbreaks and safeguard the health and well-being of the university community.

Only 71.9% of people in Arizona received one dose, 60.5% received two doses, and 40.1 % had a booster or additional dose as of Oct 15, 2022. ^21^ By comparison, in the ASU community, 94.7% of participants self-reported at least one dose, 88.9% of participants had completed their vaccination, and 62.9% of participants had a booster of a COVID vaccine. We believe that the high vaccination rate in the ASU community contributed to the low active COVID positivity rate of 0.4% based on saliva qPCR on the day of sample collection during the serosurvey study.

Similar to the earlier serosurvey^13^, two independent assays used to measure anti RBD and anti- NC antibodies showed remarkable agreement and positive correlation. This robust agreement between the assays provides further confidence in the accuracy and validity of our serological findings (Figure 1). Anti-RBD remained detectable for a duration of 6-8 months after the second dose of the vaccine.^22–24^ Moreover, our findings within the ASU community (Figure 2B) indicate that anti-RBD antibodies were still detectable even one year after vaccination. This extended duration of detectability could be attributed to the administration of booster doses received by some participants (66.9%) after completing the standard two-dose regimen. We also observed similar results in this study, where participants with breakthrough infections after vaccination exhibited higher levels of anti-RBD antibodies compared to those who had infections preceding vaccination (Figure 2C), consistent with our previous serosurvey^13^ and other research groups.^25 26^

Seroconversion was associated with the number of days after symptom onset, the severity of the disease, and the presence of co-morbidities. ^27^ However, other studies have shown that some individuals may not develop antibodies after infection and the rates of seroconversion vary. A seroprevalence study conducted in New York reported that 20% of individuals with a positive RT-PCR test result did not exhibit seroconversion.^28^ In contrast, a multicenter study conducted in Israel revealed that 5% of participants tested positive for COVID-19 through nasal swab specimens but did not show seroconversion. ^29^ Similarly, a study conducted in Alabama found that 36% of confirmed COVID-19 contacts failed to develop antibodies. ^30^ In our study, we also observed some participants remained seronegative after infection. Eighty-one participants (15.3%) that reported a history of COVID infection tested negative by both assays, some of them potentially due to the recent infection (< 2 months) or antibody decay since their SARS-CoV-2 exposure (>7 months), based on our MSD data (Figure 3A) and other studies ^31 32^. Notably, 7.4% of the 81 participants (n=6) had infection 2-6 months before collection dates, indicating that they either did not develop antibodies or produced only a small amount of antibodies, possibly due to asymptomatic infection (Figure 3B).

The Omicron variant has been associated with milder symptoms and a higher number of asymptomatic carriers, leading to increased transmission when compared to the Delta variant.^10 12 33^ Furthermore, studies have shown that the binding affinity of Omicron RBD to ACE2 is slightly weaker than that of the Beta and Delta variants. ^34 35^ Moreover, a significant proportion of individuals infected with the Omicron variant were unaware of their infection. ^36^ These findings align with the results obtained from our two serosurveys conducted during the Delta and Omicron variant waves. We observed an approximately two-fold increase in the rate of previous and unknown infections from Serosurvey I to Serosurvey II (Supplementary Table 7). Increase in the number of people with asymptomatic infection and vaccination in Serosurvey II may also explain why the median level of anti-RBD among participants 3 months after vaccination from Serosurvey II was significantly higher compared to those from Serosurvey I (Figure 2A). The observed increase in anti-RBD levels could be attributed to undetected Omicron infections, as evident in the first column of Figure 4. Notably, some longitudinal participants showed an increase in anti-RBD antibody levels over 10-fold despite reporting no change in their infection or vaccination status after Serosurvey I. Additionally, the median level of anti-RBD antibodies against Omicron subtypes was lower compared to antibodies against other variants (Supplementary Figure 1).

The most interesting finding in this study was that we detected antibodies against both NC and RBD proteins in over 1000 saliva samples, with a strong positive correlation with their corresponding serum samples. However, we observed a lower correlation for NC compared to the correlation observed for RBD (r=0.89 vs r=0.78), which could contribute to the testing of different subtypes in both assays. Typically, antibody levels measured in saliva are approximately 10-100 times lower than those found in blood samples. ^37–39^ However, in our investigation, the antibody level in saliva was determined to be 100-100,000 times lower than that detected in the blood (Figure 5). It is possible that we had higher than anticipated serum levels because of the use of the highly sensitive MDS assay in our study compared to other less sensitive serum assays employed by other laboratories. Furthermore, we identified certain saliva samples that yielded positive results while the corresponding serum samples were negative. This could potentially be due to the quality (viscosity or stickiness) of these saliva samples, potential false-positive results, also resulted in lower than expected NPA compared to DXi and MSD-platform based assays.

## Data Availability

All data produced in the present study are available upon reasonable request to the authors

## Author Contributions

JL, and VM initiated the study and design. MM, and VM developed the design. CH and VM contributed project administration, supervision, and analysis. CH, GT, VB, BB, DR, and SV designed and conducted the experiments. SW and YC performed statistical analysis. CH, VM, JL, SW, and YC wrote and revised the manuscript. All authors reviewed and approved the final version of the manuscript.

## Data sharing statement

Data may be available upon reasonable request. Contact information: Vel Murugan, Ph.D., Virginia G. Piper Center for Personalized Diagnostics, Biodesign Institute, Arizona State University, Tempe, AZ, USA

## Ethical approval

The study was approved by ASU’s institutional Review Board (IRB)(STUDY00015522).

## Participant Consent

All participants are 18+ years old and consented to participating in the study and were willing to provide their samples for the research.

## Acknowledgements

We would like to thank ASU EHS team for their logistical support and IBC oversight, communication team for their support in communicating study materials to ASU community, members of our clinical coordination team Keana Nguyen, Jerome Woodfin, Will Taylor, Abriana Gonzales, Michello Do, Kajol Majhail, Izamar Garcia, Eleni Katergaris, and Nicolas Mayhew for their help in sample accessioning, and Kylee Taylor, Bradley Bobbett, Ankita George, Scarlett Goins-Heisler, Harrison Bell, Andrew Garcia, Guillermo Trivino, and Erandi Kapuruge for their help in processing the blood tubes during the day of the serosurvey. We thank Steven Winn, and Jessica Lukosus for their help in managing the event and procurement of supplies. Funding provided by Arizona State University Knowledge Enterprise.

