## Supplementary Tables and figure for "Estimating seroprevalence of SARS-CoV-2 infection after a highly contagious Omicron outbreak: A cross sectional study in a university setting"

**Supplementary Table 1:** Anti-RBD and Anti-NC antibody seroprevalence status of the population from Serosurvey II

| Antigen Detected | Ab Sub-Type | Manufacturer | Name of the test | Assay type | Positives | Negative | Inconclusive |
|---|---|---|---|---|---|---|---|
| RBD | IgG | Beckman Coulter | Access SARS-CoV-2 IgG II | Chemiluminescent Immunoassay (CLA) | 1346 (96.3%) | 51 (3.7%) | 0 (0%) |
| | | MSD | V-PLEX COVID-19 Respiratory Panel 3 Kit | Electrochemiluminescent immunoassay (ECL) | 1369 (98%) | 28 (2%) | 0 (0%) |
| | IgM | Beckman Coulter | Access SARS-CoV-2 IgM | Chemiluminescent Immunoassay (CLA) | 28 (2%) | 1369 (98%) | 0 (0%) |
| NC | IgG | MSD | V-PLEX COVID-19 Respiratory Panel 3 Kit | Electrochemiluminescent immunoassay (ECL) | 578 (41.4%) | 819 (58.6%) | 0 (0%) |
| | Total Ab | Bio-Rad | Platelia SARS-CoV-2 Total Ab Assay | Enzyme-Linked Immunosorbent Assay (ELISA) | 546 (39.1%) | 823 (58.9%) | 28 (2%) |

**Supplementary Table 2:** Cohort characteristics and serological positive results by different assays from Serosurvey II

| Cohort | | | Spike Protein | | Nucleocapsid Protein | |
|---|---|---|---|---|---|---|
| | | | IgG | | Total ab | IgG |
| **Infection** | **Vaccine** | **n** | **Beckman** | **MSD** | **ELISA** | **MSD** |
| Yes (n=529) | YES$^{A}$ | 491 | 488 (99.4%) | 484 (98.6%) | 348 (70.9%) | 352 (71.7%) |
| | YES$^{B}$ | 4 | 4 (100%) | 3 (75%) | 4 (100%) | 4 (100%) |
| | YES$^{*}$ | 3 | 3 (100%) | 3 (100%) | 3 (100%) | 3 (100%) |
| | NO | 30 | 21 (70%) | 24 (80%) | 25 (83.3%) | 25 (83.3%) |
| | NA | 1 | 1 (100%) | 1 (100%) | 1 (100%) | 1 (100%) |
| No (n=853) | YES$^{A}$ | 805 | 781 (97%) | 799 (99.3%) | 133 (16.5%) | 160 (19.9%) |
| | YES$^{B}$ | 12 | 11 (91.7%) | 10 (83.3%) | 10 (83.3%) | 10 (83.3%) |
| | YES$^{*}$ | 3 | 3 (100%) | 3 (100%) | 2 (66.7%) | 2 (66.7%) |
| | NO | 31 | 18 (58.1%) | 26 (83.9%) | 12 (38.7%) | 13 (41.9%) |
| | NA | 2 | 1 (50%) | 1 (50%) | 0 | 0 |
| NA (n=15) | YES | 5 | 5 (100%) | 5 (100%) | 2 (40%) | 2 (40%) |
| | NA | 10 | 10 (100%) | 10 (100%) | 6 (60%) | 6 (60%) |
| Total | | 1397 | 1346 (96.3%) | 1369 (98%) | 546 (39.1%) | 578 (41.4%) |

A: Pfizer, Moderna, AstraZeneca, and Janssen; B: Covaxin, Sinopharm, *: Participants did not provide vaccine source

**Supplementary Table 3:** Seroconversion by race, age, gender, employment status, and the types of vaccines from Serosurvey II (Bold indicates statistically significant differences)

| Variable | Comparison | Anti-RBD Antibody (Access SARS-CoV-2 IgG II) | | | | Anti-NC antibody (Platelia NC total Ab) | | | |
|---|---|---|---|---|---|---|---|---|---|
| | | n | OR | 95% CI | P-value | n | OR | 95% CI | P-value |
| Race | White vs Other | 569 vs 289 | 0.98 | (0.26, 3.08) | 0.97 | 218 vs 147 | 0.55 | (0.33, 0.91) | **0.02** |
| | Asian vs Other | 355 vs 289 | 0.58 | (0.15, 1.86) | 0.38 | 144 vs 147 | 1.11 | (0.60, 2.06) | 0.73 |
| | White vs Asian | 569 vs 355 | 1.69 | (0.59, 4.89) | 0.33 | 218 vs 144 | 0.50 | (0.28, 0.88) | **0.02** |
| Age | 20-30 vs <20 | 584 vs 162 | 0.31 | (0.02, 1.58) | 0.26 | 264 vs 71 | 0.73 | (0.36, 1.40) | 0.36 |
| | 30-40 vs <20 | 187 vs 162 | 0.40 | (0.02, 3.74) | 0.46 | 79 vs 71 | 0.44 | (0.18, 1.02) | 0.06 |
| | 40-50 vs <20 | 123 vs 162 | 0.14 | (0.01, 1.24) | 0.12 | 37 vs 71 | 1.08 | (0.34, 3.51) | 0.90 |
| | 50>= vs <20 | 157 vs 162 | 0.50 | (0.02, 6.84) | 0.62 | 58 vs 71 | 0.81 | (0.28, 2.30) | 0.70 |
| | 30-40 vs 20-30 | 187 vs 584 | 1.31 | (0.34, 6.59) | 0.72 | 79 vs 264 | 0.60 | (0.31, 1.15) | 0.12 |
| | 40-50 vs 20-30 | 123 vs 584 | 0.46 | (0.10, 2.02) | 0.30 | 37 vs 264 | 1.47 | (0.56, 4.15) | 0.44 |
| | 50>= vs 20-30 | 157 vs 584 | 1.64 | (0.27, 13.17) | 0.60 | 58 vs 264 | 1.11 | (0.48, 2.63) | 0.81 |
| | 40-50 vs 30-40 | 123 vs 187 | 0.35 | (0.07, 1.52) | 0.17 | 37 vs 79 | 2.47 | (1.00, 6.60) | 0.06 |
| | 50>= vs 30-40 | 157 vs 187 | 1.25 | (0.19, 9.94) | 0.81 | 58 vs 79 | 1.86 | (0.86, 4.11) | 0.12 |
| | 50>= vs 40-50 | 157 vs 123 | 3.60 | (0.75, 25.54) | 0.13 | 58 vs 37 | 0.76 | (0.27, 1.99) | 0.58 |
| Gender | Male vs Female | 509 vs 704 | 0.78 | (0.33, 1.85) | 0.57 | 216 vs 293 | 0.83 | (0.54, 1.27) | 0.39 |
| Employment Status | Student vs Employee | 728 vs 485 | 1.84 | (0.47, 6.50) | 0.36 | 325 vs 184 | 0.94 | (0.49, 1.77) | 0.84 |
| Vaccine Group# | mRNA vaccine vs Other vaccine | 1020 vs 193 | 3.35 | (1.20, 9.29) | **0.02** | 381 vs 95 | 0.58 | (0.30, 1.07) | 0.09 |
| | Unvaccinated vs Other vaccine | N/A | N/A | N/A | N/A | 33 vs 95 | 1.43 | (0.48, 4.86) | 0.54 |
| | mRNA vaccine vs Unvaccinated | N/A | N/A | N/A | N/A | 381 vs 33 | 0.40 | (0.13, 1.01) | 0.07 |

#Unvaccinated samples were not included for the anti-RBD Antibody (Access SARS-CoV-2 IgG II)

**Supplementary Table 4:** Anti-RBD antibody difference by race, age, gender, employment status, and the types of vaccines controlling for days post vaccination from Serosurvey II (Bold indicates statistically significant differences)

| Variable | Comparison | n | beta | 95% CI | P-value |
|---|---|---|---|---|---|
| Race | White vs Other | 569 vs 289 | -25.04 | (-62.28, 12.20) | 0.19 |
| | Asian vs Other | 355 vs 289 | -7.37 | (-48.57, 33.82) | 0.73 |
| | White vs Asian | 569 vs 355 | -17.66 | (-55.56, 20.23) | 0.36 |
| Age | 20-30 vs <20 | 584 vs 162 | -10.65 | (-55.61, 34.30) | 0.64 |
| | 30-40 vs <20 | 187 vs 162 | -36.71 | (-97.60, 24.19) | 0.24 |
| | 40-50 vs <20 | 123 vs 162 | -54.95 | (-127.47, 17.57) | 0.14 |
| | 50>= vs <20 | 157 vs 162 | 30.89 | (-40.55, 102.34) | 0.40 |
| | 30-40 vs 20-30 | 187 vs 584 | -26.05 | (-73.81, 21.71) | 0.29 |
| | 40-50 vs 20-30 | 123 vs 584 | -44.30 | (-105.21, 16.62) | 0.15 |
| | 50>= vs 20-30 | 157 vs 584 | 41.55 | (-17.52, 100.61) | 0.17 |
| | 40-50 vs 30-40 | 123 vs 187 | -18.24 | (-77.34, 40.85) | 0.55 |
| | 50>= vs 30-40 | 157 vs 187 | 67.60 | (11.64, 123.56) | **0.02** |
| | 50>= vs 40-50 | 157 vs 123 | 85.85 | (25.98, 145.71) | **<0.01** |
| Sex | Male vs Female | 509 vs 704 | 18.02 | (-12.16, 48.20) | 0.24 |
| Employment Status | Student vs Employee | 728 vs 485 | 0.41 | (-44.91, 45.72) | 0.99 |
| Vaccine Group | mRNA vaccine vs Other | 1020 vs 193 | 129.71 | (86.79, 172.63) | **<0.001** |

**Supplementary Table 5:** Seroconversion by race, age, gender, employment status, and the types of vaccines from Serosurvey I and II (Bold indicates statistically significant differences)

| Variable | Comparison | Anti- RBD Antibody (Access SARS-CoV-2 IgG II | | | | Anti- NC antibody (Platelia NC total Ab) | | | |
|---|---|---|---|---|---|---|---|---|---|
| | | n | OR | 95% CI | P-value | n | OR | 95% CI | P-value |
| Race | White vs Other | 855 vs 388 | 0.73 | (0.34, 1.47) | 0.40 | 291 vs 167 | 0.53 | (0.34, 0.81) | **<0.01** |
| | Asian vs Other | 495 vs 388 | 0.77 | (0.35, 1.62) | 0.51 | 156 vs 167 | 1.31 | (0.76, 2.25) | 0.33 |
| | White vs Asian | 855 vs 495 | 0.94 | (0.51, 1.74) | 0.85 | 291 vs 156 | 0.40 | (0.24, 0.66) | **<0.001** |
| Age | 20-30 vs <20 | 854 vs 343 | 1.25 | (0.63, 2.35) | 0.51 | 324 vs 102 | 0.78 | (0.46, 1.28) | 0.33 |
| | 30-40 vs <20 | 226 vs 343 | 0.92 | (0.34, 2.70) | 0.87 | 83 vs 102 | 0.51 | (0.24, 1.04) | 0.07 |
| | 40-50 vs <20 | 144 vs 343 | 0.36 | (0.11, 1.31) | 0.11 | 44 vs 102 | 1.00 | (0.39, 2.67) | 1.00 |
| | 50>= vs <20 | 171 vs 343 | 0.87 | (0.21, 4.00) | 0.85 | 61 vs 102 | 0.94 | (0.39, 2.29) | 0.89 |
| | 30-40 vs 20-30 | 226 vs 854 | 0.74 | (0.31, 2.00) | 0.52 | 83 vs 324 | 0.65 | (0.36, 1.18) | 0.16 |
| | 40-50 vs 20-30 | 144 vs 854 | 0.29 | (0.09, 0.97) | 0.04 | 44 vs 324 | 1.29 | (0.56, 3.13) | 0.56 |
| | 50>= vs 20-30 | 171 vs 854 | 0.70 | (0.18, 3.01) | 0.61 | 61 vs 324 | 1.21 | (0.56, 2.65) | 0.63 |
| | 40-50 vs 30-40 | 144 vs 226 | 0.40 | (0.12, 1.29) | 0.12 | 44 vs 83 | 1.98 | (0.87, 4.72) | 0.11 |
| | 50>= vs 30-40 | 171 vs 226 | 0.94 | (0.24, 3.98) | 0.93 | 61 vs 83 | 1.85 | (0.89, 3.96) | 0.11 |
| | 50>= vs 40-50 | 171 vs 144 | 2.38 | (0.69, 9.37) | 0.18 | 61 vs 44 | 0.94 | (0.38, 2.27) | 0.89 |
| Gender | Male vs Female | 759 vs 979 | 0.90 | (0.53, 1.52) | 0.70 | 274 vs 340 | 0.93 | (0.64, 1.34) | 0.69 |
| Employment Status | Student vs Employee | 1209 vs 529 | 0.75 | (0.28, 1.88) | 0.55 | 422 vs 192 | 0.76 | (0.43, 1.33) | 0.34 |
| Vaccine Group# | mRNA vaccine vs Other | 1465 vs 273 | 8.11 | (4.60, 14.56) | **<0.001** | 455 vs 115 | 1.03 | (0.62, 1.69) | 0.89 |
| | Unvaccinated vs Other vaccine | N/A | N/A | N/A | N/A | 44 vs 115 | 1.81 | (0.79, 4.40) | 0.17 |
| | mRNA vaccine vs Unvaccinated | N/A | N/A | N/A | N/A | 455 vs 44 | 0.57 | (0.26, 1.17) | 0.14 |

#Unvaccinated samples were not included for the anti-RBD Antibody (Access SARS-CoV-2 IgG II)

**Supplementary Table 6:** Anti-RBD antibody difference by race, age, gender, employment status, and the types of vaccines controlling for days post vaccination from Serosurvey I and II (Bold indicates statistically significant differences)

| Variable | Comparison | n | beta | 95% CI | P-value |
|---|---|---|---|---|---|
| Race | White vs Other | 855 vs 388 | -34.22 | (-60.90, -7.53) | **0.01** |
| | Asian vs Other | 495 vs 388 | -20.37 | (-50.70, 9.95) | 0.19 |
| | White vs Asian | 855 vs 495 | -13.84 | (-40.26, 12.57) | 0.30 |
| Age | 20-30 vs <20 | 854 vs 343 | -13.70 | (-41.88, 14.48) | 0.34 |
| | 30-40 vs <20 | 226 vs 343 | -37.13 | (-79.31, 5.05) | 0.08 |
| | 40-50 vs <20 | 144 vs 343 | -64.39 | (-116.87, -11.92) | **0.02** |
| | 50>= vs <20 | 171 vs 343 | 18.61 | ( -33.51, 70.73) | 0.48 |
| | 30-40 vs 20-30 | 226 vs 854 | -23.43 | (-59.59, 12.72) | 0.20 |
| | 40-50 vs 20-30 | 144 vs 854 | -50.70 | (-97.90, -3.50) | **0.04** |
| | 50>= vs 20-30 | 171 vs 854 | 32.31 | (-14.11, 78.72) | 0.17 |
| | 40-50 vs 30-40 | 144 vs 226 | -27.26 | (-74.31, 19.78) | 0.26 |
| | 50>= vs 30-40 | 171 vs 226 | 55.74 | (10.38, 101.10) | **0.02** |
| | 50>= vs 40-50 | 171 vs 144 | 83.00 | (34.38, 131.63) | **<0.001** |
| Sex | Male vs Female | 759 vs 979 | 17.28 | (-4.38, 38.93) | 0.12 |
| Employment Status | Student vs Employee | 1209 vs 529 | -17.25 | (-52.72, 18.21) | 0.34 |
| Vaccine Group | mRNA vaccine vs Other | 1465 vs 273 | 97.24 | (67.06, 127.42) | **<0.001** |

**Supplementary Table 7:** The comparison of key outcome variables between the two serosurveys

| **Serosurvey** | **I** | **II** |
|---|---|---|
| Study Dates | 9/13-9/17, 2021 | 3/1-3/3,2022 |
| % Vaccinated (self-reported) | 91.9% | 94.7% |
| % Infected (self-reported) | 19.3% | 37.9% |
| % of positive result of anti-RBD IgG | 88.2% (Beckman) | 96.3% (Beckman) |
| | 97% (MSD) | 98% (MSD) |
| % of positive result of anti-N | 19.7% (Bio-Rad) | 39.1% (Bio-Rad) |
| | 16.1% (MSD) | 41.4 % (MSD) |
| % Unknow infection | 8.9% (Bio-Rad) | 16.5% (Bio-Rad) |
| | 9.4% (MSD) | 19.9% (MSD) |

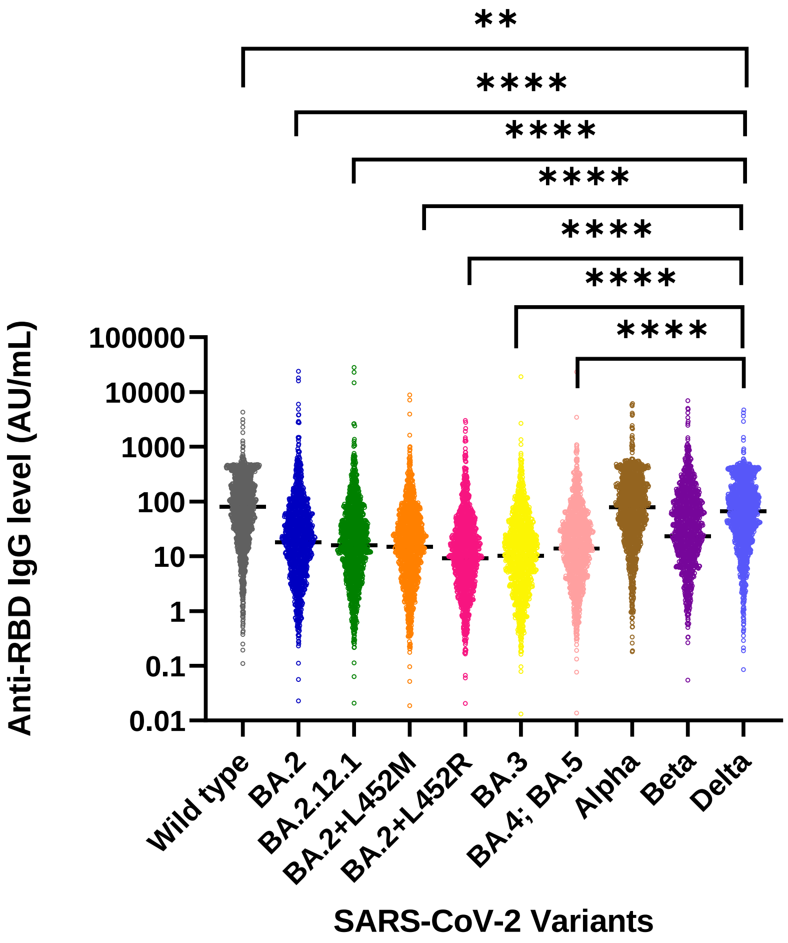


**Supplementary Figure 1: Median level of antibody levels of different variants using MSD assay.** Anti-RBD antibodies were measured using different variants of antigens with the MSD platform. *P value is calculated by the Mann-Whitney test. *p-value <0.05, **p-value <0.01, ***p-value <0.001 and ****p-value <0.0001.
